# Deciphering the Genomic Architecture of Three Major Cancers in African-Ancestry Populations

**DOI:** 10.64898/2025.12.19.25342629

**Authors:** Enoma David, Anthony Micheal Idedia, Christogonus Chichebe Ekenwaneze, Omoremime Elizabeth Dania, Olubanke Olujoke Ogunlana

**Affiliations:** Department of Biochemistry and Molecular Biology, University of Calgary, Calgary, AB, Canada; Department of Computer and Information Sciences, College of Science and Technology, Covenant University, Ota, Nigeria; Department of Biochemistry, College of Science and Technology, Covenant University, Ota, Nigeria; Covenant Applied Informatics and Communication Africa Centre of Excellence (CApIC-ACE), Covenant University, Ota, Nigeria

## Abstract

Genomic studies of cancer risk have disproportionately focused on populations of European ancestry, limiting biological insight and risk prediction in African-ancestry populations that experience a high burden of disease. Here, we analysed breast, colorectal, and prostate cancers in African-ancestry participants from the UK Biobank using ancestry-aware genome-wide association studies (GWAS), SNP-based heritability estimation, fine-mapping, transcriptome-wide association studies (TWAS), and polygenic risk scoring (PRS). SNP-based heritability analyses revealed a comparatively high point estimate of common-variant heritability for colorectal cancer risk in African-ancestry individuals, alongside more modest estimates for breast and prostate cancer. Five loci reached genome-wide significance (*p* < 5×10−□), including four colorectal cancer loci (notably rs111448231 in *RYR2*) and one novel breast cancer locus (rs78768133). Gene-based burden testing identified eight prostate cancer-associated genes (*MRPL45, PSMD8, GGN, SPRED3, FAM98C, BCLAF1, MTFR2, and NELL2*) with FDR-significant associations, clustering within biologically plausible chromosomal regions on chr19q13 and chr6q23. Transcriptome-wide association analysis identified *CYTH2 (ENSG00000105443.13)* as a significant gene for prostate cancer. Polygenic risk scores incorporating African-ancestry linkage disequilibrium demonstrated heterogeneous predictive performance across cancers, with modest discrimination for colorectal and breast cancer and substantially stronger performance for prostate cancer (AUC = 0.89). Together, these findings delineate ancestry-relevant cancer genetic architectures and demonstrate the importance of population-matched genomic approaches for equitable precision oncology.

## Introduction

Cancer is a leading cause of mortality worldwide, with prostate, breast, and colorectal cancers accounting for a substantial number of the global disease burden. In 2022, the International Agency for Research on Cancer (IARC) estimated about 20 million new cancer cases and nearly 10 million cancer deaths globally, with colorectal cancer causing approximately 900,000 deaths (9.3%), breast cancer 670,000 deaths (6.9%), and prostate cancer remaining a major contributor to cancer mortality ^1,2^. The global cancer burden is predicted to escalate substantially by 2050, driven by population growth, ageing, and shifts in risk factors, with the sharpest increases projected in low- and middle-income countries, including large parts of Africa ^3^.

Prostate cancer (PCa) remains the most frequently diagnosed cancer in men, making up nearly 30% of all male cancer cases in 2025 ^4^.

PCa is a significant health concern among men of African ancestry (AA), who experience the highest global incidence and mortality rates for this disease ^5,6^. The lifetime risk is about 1 in 6, and they experience the highest incidence rate of any ethnic group, around 191.5 cases per 100,000. PCa mortalities among AA men are roughly 36.9 per 100,000, almost double the rate seen in other groups and consistently higher than those observed in European Ancestry (EA) males across all age ranges ^4^. The variation in incidence and outcome is attributed to both genetic and non-genetic factors, including ancestry-specific risk loci, environmental exposures, access to healthcare, and social determinants ^7,8^.

Large multi-country GWAS in African populations, including Ghana, Nigeria, Senegal, South Africa, and Uganda have provided important insights into the genomic landscape of prostate cancer (3,963 cases and 3,509 controls) ^9,10^. These studies show substantial heterogeneity in effect sizes across African populations, reflecting diverse population histories and evolutionary pressures ^10^. Notably, germline mutations in genes such as *BRCA2* significantly elevate the risk of aggressive disease, while several risk single-nucleotide polymorphisms (SNPs) rare in Europeans are common and functionally relevant in African populations. For example, a *KLF5* driver mutation and distinct molecular subtypes have been identified in southern African prostate tumors ^10,11^. Environmental, socioeconomic, and healthcare-related determinants further compound biological risk, leading to later-stage diagnoses and more aggressive disease phenotypes ^12,13^. These findings underscore the need for integrative approaches that combine genomics and epidemiology to improve early detection and outcomes among AA men ^14^.

Breast cancer (BCa) is globally prevalent, but woman of AA often display more aggressive tumor subtypes and lower survival rates ^15^. A landmark genome-wide association study involving 18,034 cases and 22,104 controls of AA identified twelve loci significantly associated with breast cancer risk, including variants strongly linked to the aggressive triple-negative breast cancer (TNBC) subtype. Approximately 8% of AA women carry a combination of six high-risk variants that collectively increase their likelihood of TNBC by more than fourfold ^16^. Inherited pathogenic variants in *BRCA1* and *BRCA2* also occur at higher frequencies in several African populations, including Algerian, Moroccan, Nigerian, and Egyptian groups relative to global averages ^17^. Genetic ancestry has been shown to influence the tumor immune microenvironment, with higher African genomic ancestry associated with distinct immune response pathways and poorer prognosis subtypes such as HER2+ and TNBC ^18^.

Colorectal cancer (CRC) shows significant disparities across ancestral groups, with individuals of AA experiencing higher incidence and mortality rates compared to those of European ancestry (EA) ^19^. In a comprehensive genomic analysis of more than 46,000 colorectal adenocarcinoma cases, AA patients were significantly younger at the time of biopsy or surgery (median age 58) compared to EA patients (median age 61; Q ≤ 0.0001) ^20^. AA CRC tumors demonstrated more frequent somatic alterations in genes such as *KRAS*, *APC*, and *PIK3CA*, impacting disease onset and progression ^20^. Prior studies have reported an elevated burden of hereditary CRC syndromes and early-onset CRC among African-ancestry populations, partly attributable to unique mutation spectra and variant frequencies^21^. AA CRC cases also had a lower prevalence of microsatellite instability-high (MSI-H) tumors (3.9% vs. 5.5% in EA), with potential implications for immunotherapy responsiveness, and exhibited lower rates of hypermutated tumors ^20^. These emerging insights reinforce the urgent need for ancestry-inclusive genomic research in CRC.

The genetic architecture of major cancers in AA populations is of substantial scientific and public health significance. Disparities in cancer incidence, presentation, and survival persist between AA and European or Asian ancestry populations ^22^. These disparities are influenced by socioeconomic, environmental, and genetic factors. Yet most GWAS have focused on EA cohorts, leaving knowledge gaps and limiting accurate risk prediction, biomarker discovery, and equitable clinical translation for AA populations ^9,10,20^.

GWAS in AA populations have begun to identify cancer risk loci that may be missed or imperfectly tagged in EA focused studies, including BCa susceptibility signals ^16,23,24^, CRC loci ^21,25^, and PCa risk variants enriched in people of African ancestry ^26,27^. These studies are particularly informative because African genomes generally exhibit higher genetic diversity and shorter linkage disequilibrium (LD) blocks ^28,29^, improving localization of association signals To complement variant-level GWAS, transcriptome-wide association studies (TWAS) can aggregate cis-genetic effects into gene-level tests by associating genetically predicted expression with disease risk ^27^

Progress toward reducing cancer disparities in African-ancestry populations requires systematic efforts to identify ancestry-specific risk variants, refine disease-relevant biological pathways, and develop prediction tools tailored to diverse populations. Such efforts are crucial for advancing equitable precision oncology and ensuring that genomic medicine benefits populations that have been historically underrepresented in large-scale genetic studies.

In this study, we aimed to characterise and compare the genetic architecture of breast, colorectal, and prostate cancers in African-ancestry individuals using a unified, ancestry-aware analytical framework. Specifically, we integrated genome-wide association studies, SNP-based heritability estimation, fine-mapping, gene-based testing, transcriptome-wide association analyses, and polygenic risk prediction to identify ancestry-relevant risk loci and to evaluate the transferability and performance of genomic risk models across cancers.

## Results

### Post-Quality Control of input variants and GWAS Calibration

After applying the harmonized variant-level quality control filters the final set of SNPs retained for genome-wide analyses ranged from approximately 509,000 to 526,000 variants per cancer dataset. Specifically, breast cancer retained 526,048 SNPs, prostate cancer retained 509,066 SNPs, and colorectal cancer retained 515,671 SNPs, from an initial ∼783,866 array-genotyped markers prior to QC. Principal component analysis (PCA) ^30^ of the final genotype data for the three African-ancestry cancer cohorts (Supplementary figures 1-3) showed a single, continuous African/African-admixed cluster with modest internal dispersion along the first two principal components and no evidence of discrete sub-populations. Quantile–quantile (QQ) plots (Supplementary figures 4 – 6). of the Fast-LMM GWAS test statistics demonstrated excellent calibration in all three traits. Genomic inflation factors ^31^ (*λ_GC_*) were 1.004 for CRC, 1.018 for PCa, and 0.998 for BCa, indicating minimal residual population stratification or technical artefact after mixed-model correction.

**Figure 1.**
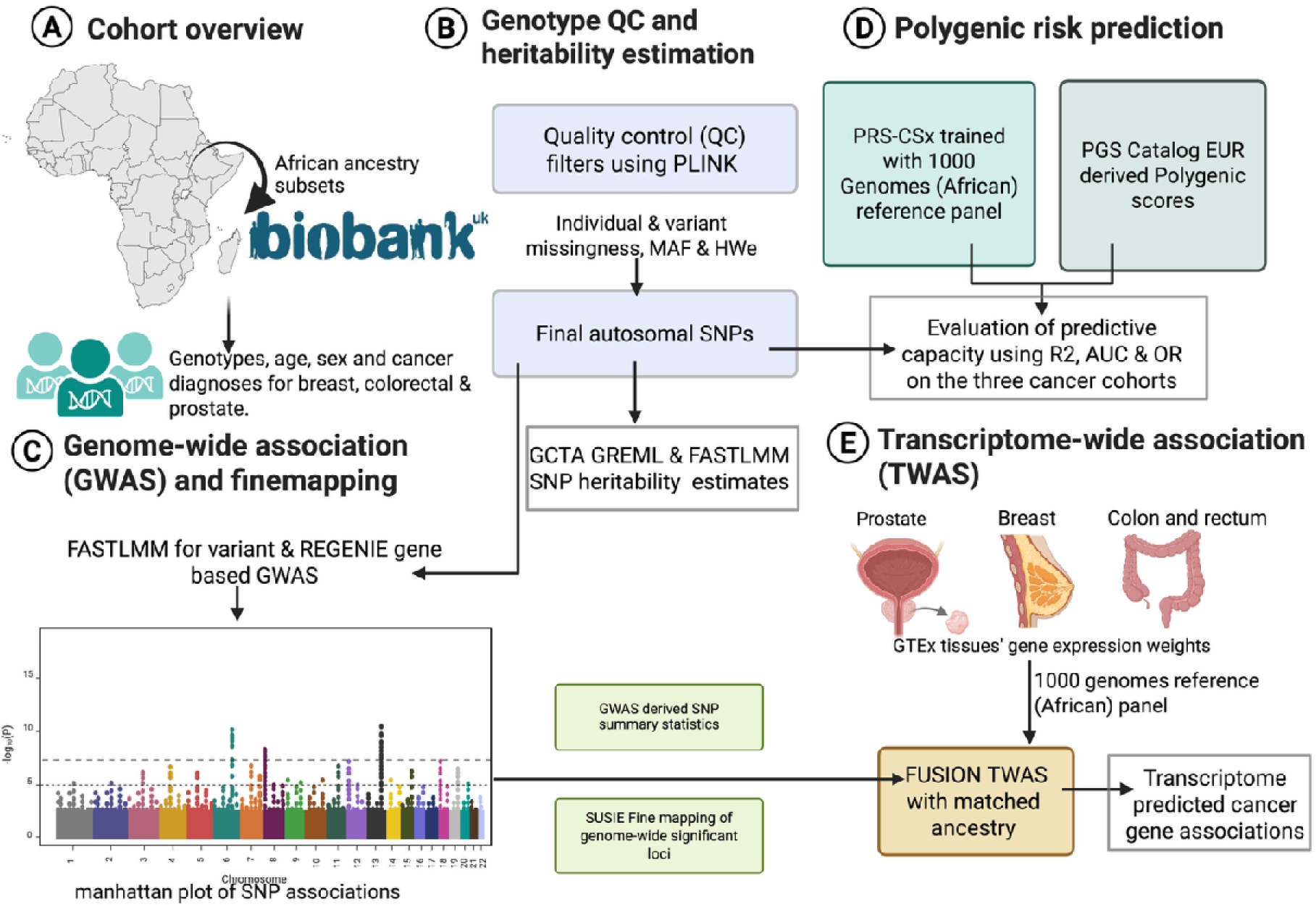
Overview of African-ancestry cancer genetics study design. (A) African-ancestry subsets of the UK Biobank were ascertained with genome-wide genotypes, age, sex, and incident or prevalent diagnoses of breast (BCa), colorectal (CRC) and prostate (PCa) cancer. (B) Genotype quality control was performed using PLINK, including individual and SNP missingness filters, minor-allele frequency and Hardy–Weinberg equilibrium thresholds, yielding a final set of autosomal variants for downstream analyses. SNP-heritability was estimated with GCTA GREML and FASTLMM. (C) Genome-wide association studies (GWAS) were run using FASTLMM for variant-level tests and REGENIE for gene-based tests. Genome-wide significant loci were then subjected to SuSiE fine-mapping using an African-ancestry LD reference panel. (D) Polygenic risk prediction was evaluated using two approaches: PRS-CSx models trained with 1000 Genomes African reference data and polygenic scores obtained from the PGS Catalog (originally derived from largely European studies). Predictive capacity for each score was assessed by odds ratio per standard deviation (OR/SD), area under the ROC curve (AUC) and liability-scale pseudo-R² in the three cancer cohorts. (E) Transcriptome-wide association studies (TWAS) were conducted using FUSION with GTEx v8 tissue-specific expression weights (prostate, breast, colon/rectum) and 1000 Genomes African LD, enabling identification of transcriptome-predicted cancer risk genes.

### SNP-Based heritability in African-Ancestry Cancers

We estimated SNP-based heritability (See Methods) for BCa, CRC, and PCa in African-ancestry UK Biobank participants using both GCTA-GREML ^32^ and FaST-LMM ^33^, which yielded highly concordant results (Table 2). For breast cancer, GCTA estimated a modest heritability of h² = 0.114 (SE = 0.146), with FaST-LMM producing an almost identical estimate (h² = 0.115). For colorectal cancer, we observed a substantially higher heritability: h² = 0.603 (SE = 0.209) from GCTA, closely matched by FaST-LMM (h² = 0.609).

This represents a strong polygenic contribution to CRC risk in African-ancestry individuals, even after stringent quality control and sample size constraints. For prostate cancer, heritability was lower but consistent across approaches, with h² = 0.081 (SE = 0.092) from GCTA and h² = 0.084 from FaST-LMM. The close agreement between GCTA and FaST-LMM across all three cancer phenotypes demonstrates the robustness of these estimates. Notably, CRC exhibited a markedly higher SNP heritability (∼0.60) relative to BCa and PCA, suggesting stronger underlying common-variant genetic architecture in this African-ancestry cohort.

### Lead SNP–Gene mappings and genome-wide significant loci

Using g:Profiler-based functional annotation ^34^ of lead SNPs (p < 1×10−□), we identified a compact set of variants mapping to biologically plausible genes for colorectal cancer (CRC), breast cancer (BCa), and prostate cancer (PCA) (Figure 2 and Table 1). Across all three cancers, five loci achieved genome-wide significance ^35^ (p < 5×10−□). These SNPs include four loci for colorectal cancer (notably rs111448231 in *RYR2*) and one novel locus for breast cancer (rs78768133). These genome-wide hits represent the most robust SNP–gene signals in the African-ancestry analyses and define high-priority candidates for follow-up functional work.

**Figure 2.**
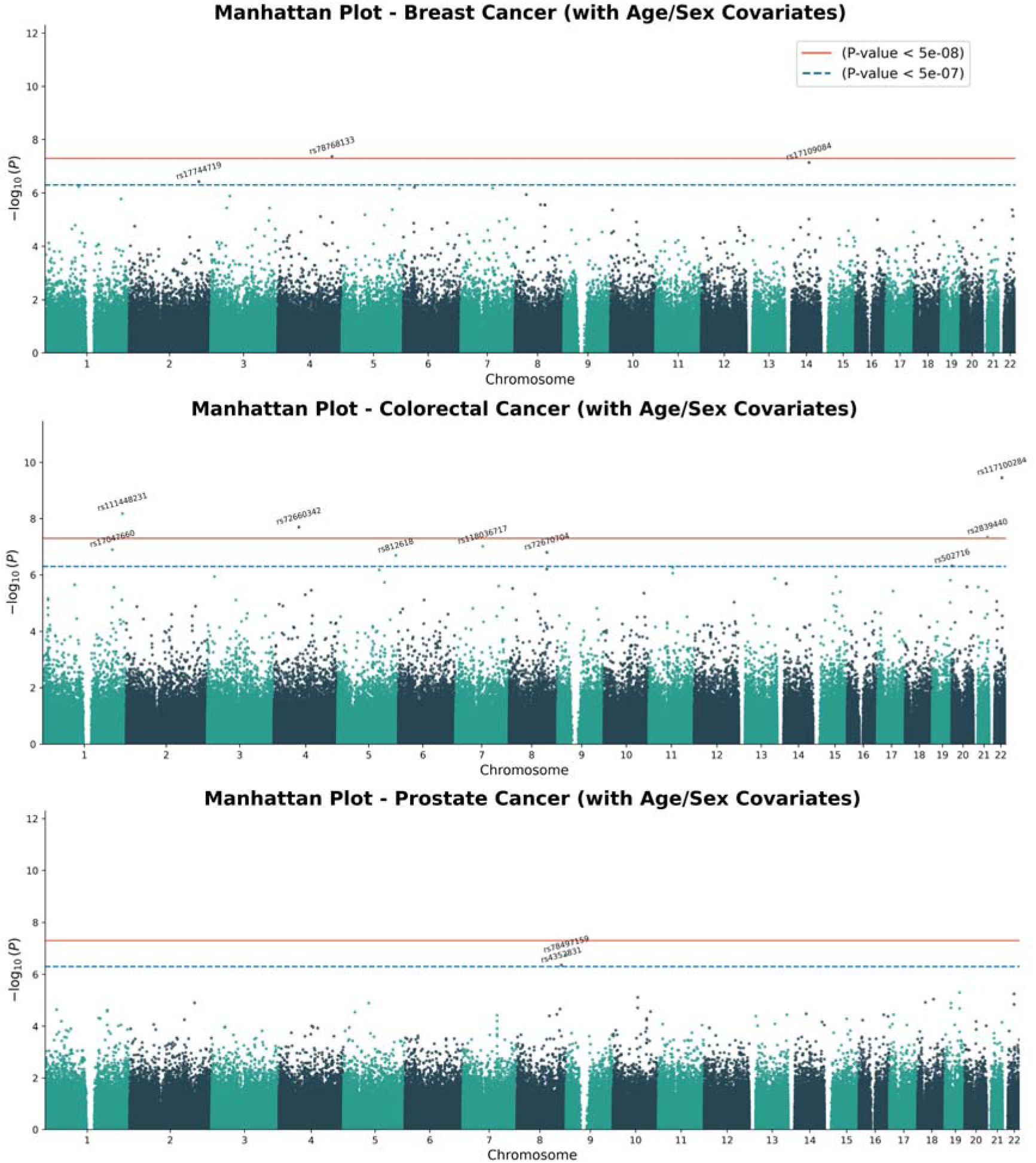
Genome-wide association results for breast, colorectal and prostate cancer in African-ancestry UK Biobank participants. Manhattan plots display –logOO(P) for SNP associations across autosomes for (top) breast cancer (BCa), (middle) colorectal cancer (CRC) and (bottom) prostate cancer (PCa), adjusted for age and sex. The red horizontal line marks the conventional genome-wide significance threshold (PO<O5×10−□), and the blue dashed line indicates a suggestive threshold (PO<O5×10−□). The most strongly associated lead variants highlighted in the plot include rs78768133 (BCa), rs117100284 and rs111448231 (CRC), and rs78497159 and rs4352831 (PCa), with full summary statistics provided in Table 1.

**Table 1.** Lead SNPs associated with breast, colorectal and prostate cancer in African-ancestry UK Biobank participants. The table lists all variants with PO<O1×10^−5^ from the FASTLMM GWAS for breast (BCa), colorectal (CRC) and prostate (PCA) cancer. For each SNP we report chromosome, closest or mapped Ensembl gene ID(s), gene symbol(s), association P-value and cancer phenotype. Notable examples include rs117100284 (chr22, CRC; P = 3.55×10^−1□^), rs111448231 mapping to RYR2 (CRC), rs17047660 in CR1 (CRC), rs78497159 in DMRT1 (PCA), and several breast-cancer signals near TFPI2, DOCK2, PFKFB3 and other genes. Loci without an assigned gene indicate variants not confidently mapped to a specific protein-coding gene.

| SNP | Chr | Gene ID | Gene Name | P-value | Cancer |
| --- | --- | --- | --- | --- | --- |
| rs117100284 | 22 |  |  | 3.55E-10 | CRC |
| rs111448231 | 1 | ENSG00000198626 | *RYR2* | 6.64E-09 | CRC |
| rs72660342 | 4 |  |  | 2.01E-08 | CRC |
| rs78768133 | 4 |  |  | 4.29E-08 | BCa |
| rs2839440 | 21 |  |  | 4.54E-08 | CRC |
| rs17109084 | 14 |  |  | 7.14E-08 | BCa |
| rs118036717 | 7 |  |  | 9.76E-08 | CRC |
| rs17047660 | 1 | ENSG00000203710 | *CR1* | 1.29E-07 | CRC |
| rs72670704 | 8 | ENSG00000164796 | *CSMD3* | 1.61E-07 | CRC |
| rs78497159 | 9 | ENSG00000137090 | *DMRT1* | 1.79E-07 | PCA |
| rs812618 | 5 |  |  | 2.05E-07 | CRC |
| rs17744719 | 2 |  |  | 3.76E-07 | BCa |
| rs4352831 | 8 |  |  | 4.39E-07 | PCA |
| rs502716 | 20 |  |  | 4.88E-07 | CRC |
| rs11233800 | 11 | ENSG00000172890 | *NADSYN1* | 5.52E-07 | CRC |
| rs79041939 | 1 |  |  | 5.84E-07 | BCa |
| rs3130605 | 6 |  |  | 6.07E-07 | BCa |
| rs72685816 | 8 | ENSG00000164796 | *CSMD3* | 6.29E-07 | CRC |
| rs73227485 | 7 | ENSG00000105825 | *TFPI2* | 6.59E-07 | BCa |
| rs72779318 | 5 |  |  | 6.71E-07 | CRC |
| rs75759774 | 5 | ENSG00000134516 | *DOCK2* | 6.90E-07 | BCa |
| rs80165586 | 11 |  |  | 8.79E-07 | CRC |
| rs72633387 | 8 |  |  | 1.16E-06 | BCa |
| rs55704627 | 3 | ENSG00000151789 | *ZNF385D* | 1.16E-06 | CRC |
| rs12437516 | 15 |  |  | 1.17E-06 | CRC |
| rs6801157 | 3 | ENSG00000187672 | *ERC2* | 1.31E-06 | BCa |
| rs72650580 | 13 | ENSG00000204442 | *NALF1* | 1.37E-06 | CRC |
| rs74387320 | 19 | ENSG00000105609 | *LILRB5* | 1.56E-06 | CRC |
| rs2489320 | 1 | ENSG00000185842 | *DNAH14* | 1.69E-06 | BCa |
| rs62383900 | 5 | ENSG00000158458 | *NRG2* | 1.85E-06 | CRC |
| rs74841697 | 14 |  |  | 2.04E-06 | CRC |
| rs17113161 | 1 | ENSG00000152078,ENSG00000271092 | *TLCD4,TLC*<br>*D4-RWDD3* | 2.23E-06 | CRC |
| rs7270676 | 20 | ENSG00000124233 | *SEMG1* | 2.61E-06 | CRC |
| rs8130742 | 21 |  |  | 2.66E-06 | CRC |
| rs12087287 | 1 |  |  | 2.74E-06 | CRC |
| rs56117219 | 8 |  |  | 2.76E-06 | BCa |
| rs75912510 | 8 |  |  | 2.82E-06 | BCa |
| rs75927026 | 8 |  |  | 2.82E-06 | BCa |
| rs17151790 | 8 | ENSG00000175806 | *MSRA* | 2.98E-06 | CRC |
| rs11729080 | 4 |  |  | 3.45E-06 | CRC |
| rs77178871 | 3 | ENSG00000160799 | *CCDC12* | 3.64E-06 | BCa |
| rs11743933<br>7 | 21 | ENSG00000185658 | *BRWD1* | 3.67E-06 | CRC |
| rs77478751 | 3 | ENSG00000169760 | *NLGN1* | 3.68E-06 | BCa |
| rs74672539 | 17 | ENSG00000158955 | *WNT9B* | 3.69E-06 | CRC |
| rs12148207 | 15 |  |  | 3.90E-06 | CRC |
| rs76179800 | 5 |  |  | 4.22E-06 | BCa |
| rs73176670 | 22 | ENSG00000100403 | *ZC3H7B* | 4.28E-06 | BCa |
| rs12359173 | 10 | ENSG00000170525 | *PFKFB3* | 4.36E-06 | BCa |
| rs11198880 | 10 | ENSG00000198873 | *GRK5* | 4.38E-06 | CRC |
| rs10459627 | 15 | ENSG00000166035 | *LIPC* | 4.53E-06 | CRC |
| rs11753419<br>7 | 8 |  |  | 4.77E-06 | CRC |
| rs80233874 | 4 |  |  | 4.95E-06 | CRC |
| rs77933005 | 19 |  |  | 5.09E-06 | PCA |
| rs365833 | 22 |  |  | 5.77E-06 | PCA |
| rs34889768 | 5 |  |  | 6.67E-06 | BCa |
| rs75656779 | 1 |  |  | 6.81E-06 | CRC |
| rs79358807 | 22 | ENSG00000186732 | *MPPED1* | 7.36E-06 | BCa |
| rs848215 | 1 |  |  | 7.52E-06 | CRC |
| rs79083223 | 1 | ENSG00000143669 | *LYST* | 7.60E-06 | CRC |
| rs6927735 | 6 |  |  | 7.65E-06 | CRC |
| rs11591051 | 22 |  |  | 7.67E-06 | BCa |
| 8 |  |  |  |  |  |
| rs75459996 | 3 | ENSG00000175161 | *CADM2* | 7.67E-06 | CRC |
| rs34284214 | 10 | ENSG00000166321 | *NUDT13* | 7.83E-06 | PCA |
| rs55905897 | 22 | ENSG00000100218,ENSG00000128266 | *RSPH14,GN*<br>*AZ* | 8.68E-06 | CRC |
| rs7296226 | 12 |  |  | 9.17E-06 | CRC |
| rs16940123 | 18 | ENSG00000078043 | *PIAS2* | 9.20E-06 | PCA |
| rs55908223 | 19 | ENSG00000130844 | *ZNF331* | 9.41E-06 | CRC |
| rs74666579 | 14 | ENSG00000100731 | *PCNX1* | 9.58E-06 | BCa |
| rs60132535 | 7 | ENSG00000189320 | *FAM180A* | 9.63E-06 | BCa |

In addition, we observed a small number of highly significant associations with 5×10−□ ≤ p < 5×10−□ ^36,37^ which did not pass the conventional genome-wide threshold but show strong statistical evidence of association and are likely to be confirmed in larger African-ancestry cohorts. Finally, the full annotation table (Table 1) includes all suggestive loci with p < 1×10−□, providing a broader catalogue of candidate SNP–gene mappings that may inform further downstream analyses.

### Finemapping of African-Ancestry cancer loci

SuSiE ^38^ fine-mapping resolved the architecture of African-ancestry cancer loci with high resolution due to the sharper LD structure characteristic of African genomes. Across all loci examined, SuSiE typically identified single, compact causal signals, reflecting limited long-range LD and increased localization precision relative to European-derived LD patterns. Prostate cancer locus at *rs117100284* (chr22:38.45 Mb) (Figure 3A). The prostate cancer locus centred on rs117100284 fine-mapped to a single 95% credible set, indicating one dominant causal component in African-ancestry individuals. After alignment of 45 variants with the AFR LD panel, the lead SNP showed the highest posterior inclusion probability, with a narrow cluster of nearby variants (AFR r² > 0.8) sharing secondary weight (Figure 3B). No additional credible sets were detected, suggesting that the regional association signal is monogenic rather than composed of multiple independent effects. PIP and −log10(P) profiles showed strong concordance, supporting the robustness of the inferred causal structure.

**Figure 3.**
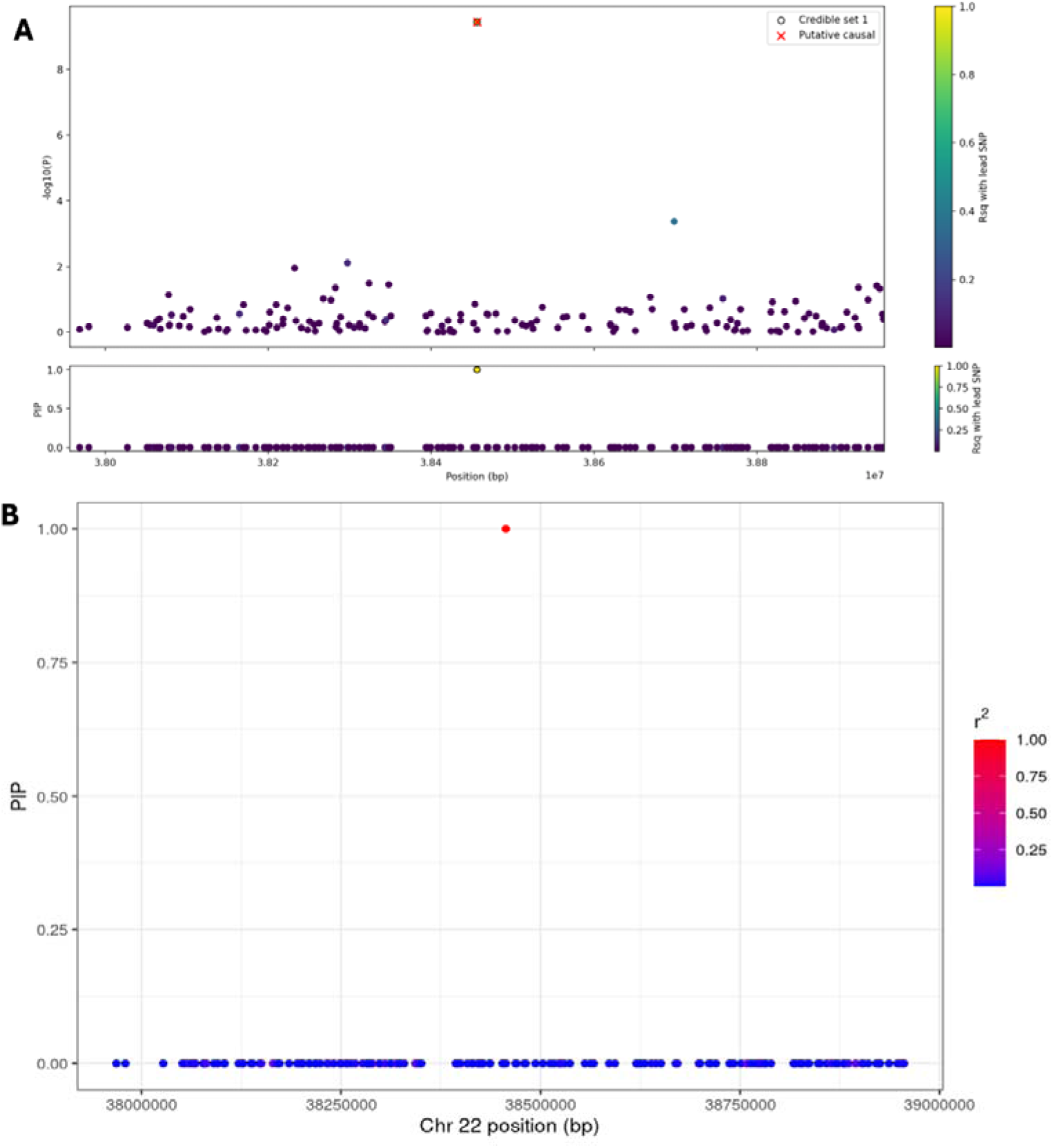
SuSiE fine-mapping of the colorectal cancer locus at chromosome 22 tagged by rs117100284. (A) Regional association plot for SNPs within ±500Okb of rs117100284 (chr22:38.46OMb) in the colorectal cancer GWAS. Points show –logOO(P) for each variant, coloured by LD (r²) with the lead SNP using African-ancestry LD from the AFR_hg38 reference panel. Variants belonging to the 95% credible set identified by SuSiE are marked (red), whereas nearby signal(s) outside the credible set are shown in other colours. (B) Posterior inclusion probabilities (PIP) from SuSiE across the same region. A single high-confidence credible set is observed, with one variant reaching PIP ≈ 1.0 and the remainder having near-zero PIP, consistent with a largely single-causal-variant architecture at this locus in the African-ancestry colorectal cancer cohort.

### Gene-based burden test highlights prostate-specific risk genes

Using REGENIE, Gene-based burden tests identified modest but trait-specific signals across the three cancers (Supplementary data 4 – 6). For breast (BCa) and colorectal cancer(CRC), none of the top-ranked genes reached genome-wide or FDR-adjusted significance; the strongest associations are shown in Supplementary Table 2. In contrast, prostate cancer (PCA) exhibited a small but coherent set of significant gene-level associations. Using the REGENIE burden test, eight genes, *MRPL45, PSMD8, GGN, SPRED3, FAM98C, BCLAF1, MTFR2,* and *NELL2* surpassed the FDR threshold (q < 0.05) (Table 3), with p-values ranging from ∼1×10−□ to 4×10−□. These genes cluster within biologically plausible chromosomal regions (notably chr19q13 and chr6q23), suggesting localized aggregation of prostate-cancer risk variants in African-ancestry individuals. No genome-wide significant signals were observed in the REGENIE analyses, though PCA again showed the strongest near-significant enrichment compared with BCa and CRC.

### Transcriptome-wide association results across three cancer cohorts

Trait-specific TWAS analyses were performed for colorectal, breast, and prostate cancer (Figure 4, Supplementary data 7-9) using GTEx v8 tissue prediction models together with African-ancestry GWAS summary statistics and ancestry-matched LD from the 1000 Genomes AFR panel (Figure 1E). After applying Bonferroni correction within each trait, only one gene reached transcriptome-wide significance: *CYTH2* (*ENSG00000105443.13*) in the prostate cancer analysis (TWAS Z = 4.15, TWAS P = 3.3 × 10−□) (Figure 4).

**Figure 4.**
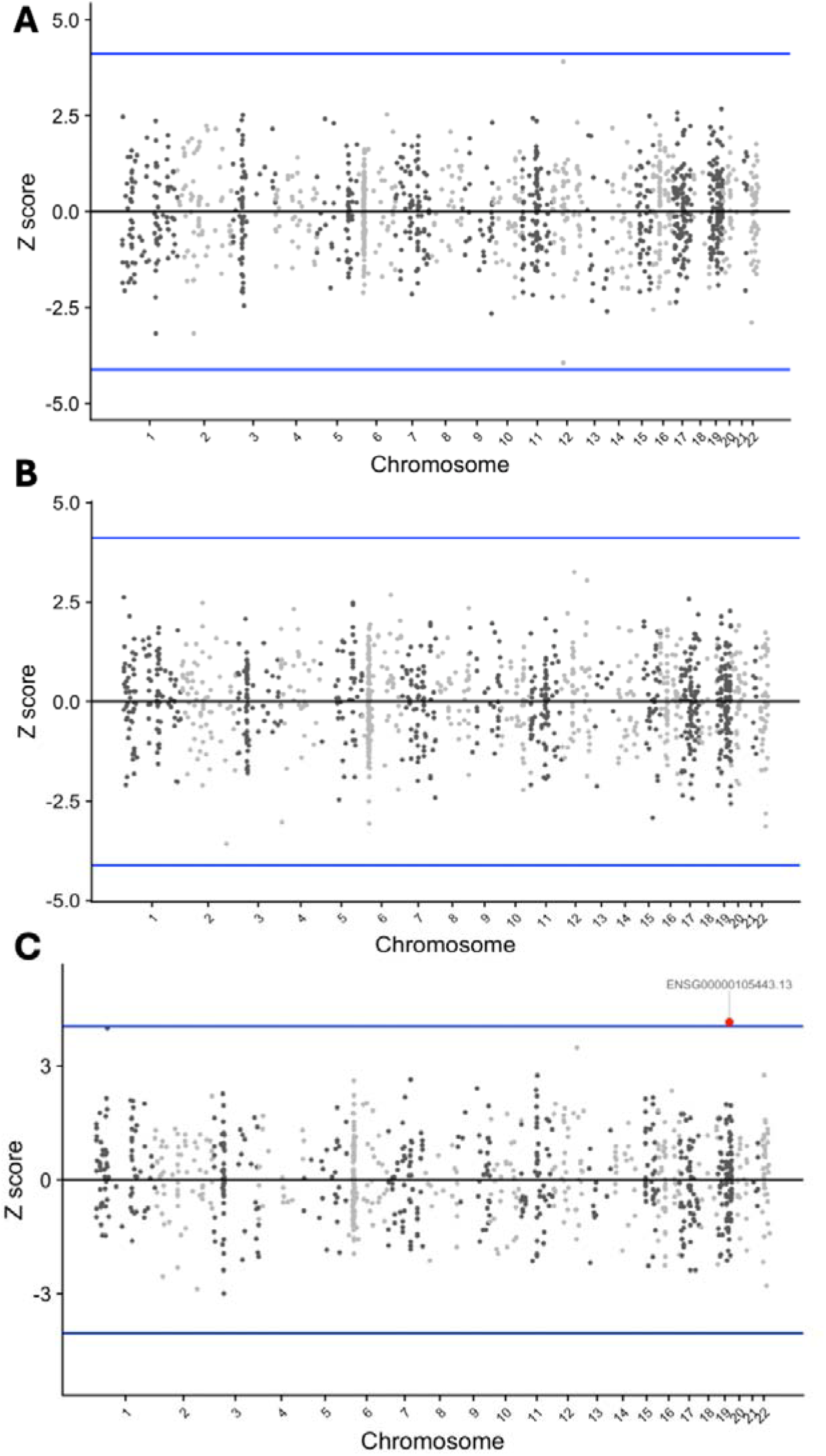
Transcriptome-wide association of breast, colorectal and prostate cancer using ancestry-matched TWAS. Z-scores from FUSION TWAS are plotted by genomic position for (A) breast, (B) colorectal and (C) prostate cancer, using GTEx v8 expression weights for the corresponding tissues and African-ancestry GWAS summary statistics. Horizontal blue lines indicate Bonferroni-corrected significance thresholds. Grey points represent all tested genes, and darker points highlight genes with stronger evidence of association. In the prostate TWAS (panel C), *CYTH2* (*ENSG00000105443.13*) emerges as the top association, with additional suggestive genes observed for breast and colorectal cancer (see Supplementary Tables).

Regional association analyses converged on a single 19q locus where both SNP- and gene-level signals implicated the *CYTH2* region (Supplementary Figure 8). A tight cluster of variants in high LD around ∼48.5 Mb showed the strongest association peak, centred on the TWAS-prioritised gene (Supplementary Figure 7). This pattern suggests that risk in this region is driven by a locally coherent block of variants at the *CYTH2* locus. A key limitation of these analyses is that SNP coverage in the UK Biobank African-ancestry cohort was restricted to directly genotyped array variants. This substantially reduces the number of available cis-regulatory SNPs compared with imputed datasets and disproportionately affects African-ancestry analyses.

### Polygenic risk score performance in African-Ancestry validation cohorts

We next evaluated the performance of two complementary PRS strategies in the African-ancestry UK Biobank cohorts, PRS-CSx ^39^ scores trained on multi-ancestry discovery GWAS with African LD reference and trait-specific scores from the PGS Catalog (Set 2; PGS000377 for CRC, PGS002241 for PCA, PGS002242 for BCa), originally derived largely in European cohorts but harmonized to GRCh38. All models were adjusted for age at recruitment and sex (See Methods).

For colorectal cancer (CRC; 112 cases, 989 controls) (Supplementary Table 1), both scores yielded modest but directionally consistent risk stratification. The PRS-CSx score produced an OR per SD of 1.19 (95% CI: 0.96–1.46; p = 0.11), with an AUC of 0.63 and a Nagelkerke ΔR² of 0.0051 beyond covariates alone. The PGS000377 score showed a similar effect (OR per SD 1.17, 95% CI: 0.96–1.42; p = 0.12), AUC of 0.63, and ΔR² of 0.0045, indicating comparable and modest incremental predictive value.

In prostate cancer (PCA; 250 cases, 2261 controls) (Supplementary Table 1), both approaches achieved strong predictive capacity. The African-tuned PRS-CSx score attained an OR per SD of 1.40 (95% CI: 1.20–1.63; p = 1.95×10−□), with an AUC of 0.89 and ΔR² of 0.0131. The PGS002241 score also performed well (OR per SD 1.30, 95% CI: 1.11–1.52; p = 1.07×10⁻³; AUC = 0.89) but explained slightly less additional variance (ΔR² = 0.0075).

For breast cancer (BCa; 166 cases, 1399 controls) (Supplementary Table 1), age and sex already conferred substantial predictive capacity (baseline AUC ≈ 0.77), and both PRS contributed only modest incremental information. The PRS-CSx score had an OR per SD of 1.11 (95% CI: 0.94–1.32; p = 0.22), AUC of 0.77, and ΔR² of 0.0018. The PGS002242 score showed a nominally stronger association (OR per SD 1.21, 95% CI: 1.02–1.44; p = 2.81×10⁻²), AUC of 0.77, and ΔR² of 0.0057, but absolute gains remained small. Together, these results indicate that PRS-CSx, which explicitly models African LD, even in small sample sizes provides slightly better calibrated risk prediction (Figure 5) than non-matched European LD with significantly larger sample sizes.

**Figure 5.**
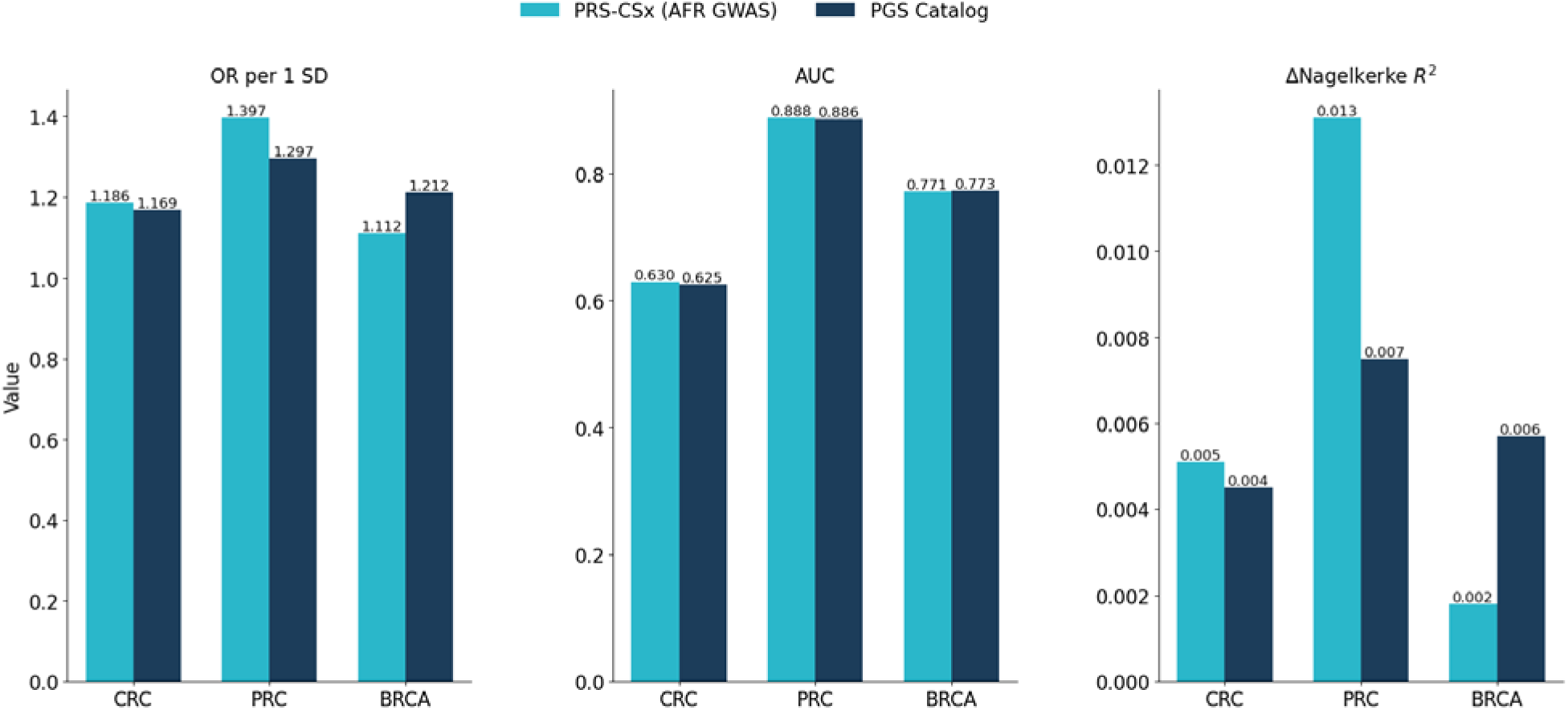
Performance of cross-ancestry and catalog polygenic scores for African-ancestry cancer risk. Bar plots compare prediction performance for colorectal (CRC), prostate (PCa) and breast (BCa) cancer between PRS-CSx scores trained using African-ancestry 1000 Genomes reference data (teal) and polygenic scores taken from the PGS Catalog (navy). Metrics shown are: (left) odds ratio per one standard deviation increase in the polygenic score (OR/SD); (middle) area under the receiver operating characteristic curve (AUC); and (right) change in liability-scale pseudo-R² relative to a covariate-only model (ΔR²).

## Discussion

In this study, we applied an ancestry-aware genomic framework (Figure 1) to dissect the genetic architecture of breast, colorectal, and prostate cancers in African-ancestry individuals from the UK Biobank. Using mixed-model GWAS with excellent calibration (λGC = 0.998–1.018), SNP-based heritability estimation, fine-mapping, gene-based testing, transcriptome-wide association studies (TWAS), and polygenic risk scoring (PRS), we identify ancestry-relevant features that differ markedly across cancer types. Together, these results highlight both shared and cancer-specific genetic architectures while underscoring the importance of population-matched genomic analyses for equitable precision oncology.

A striking observation was the substantially higher SNP-based heritability estimate for colorectal cancer (h² ≈ 0.60) relative to breast (h² ≈ 0.11) and prostate cancer (h² ≈ 0.08) (Table 2). Although confidence intervals remain wide due to limited sample size, this pattern is consistent across both GCTA-GREML and FaST-LMM implementations, indicating a robust signal rather than methodological artefact ^32,33^ (Yang et al., 2011; Lippert et al., 2011). The comparatively strong common-variant contribution to colorectal cancer risk aligns with prior evidence of pronounced germline susceptibility in African-ancestry CRC populations, including ancestry-enriched risk alleles and earlier disease onset ^20,25^

**Table 2.** SNP-heritability of breast, colorectal and prostate cancer in African-ancestry UK Biobank participants. Sample sizes used for heritability analyses (N), GCTA GREML SNP-heritability estimates 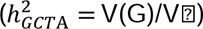, corresponding standard errors (SE), and FASTLMM-based h² estimates are shown for each cancer phenotype.

| Trait | N(heritability analysis) | $h^2_{GCTA}$<br>($V(G)/V(\epsilon)$) | SE ($h^2_{GCTA}$) | $h^2_{FaST-LMM}$ |
| --- | --- | --- | --- | --- |
| BCa | 1,210 | 0.114 | 0.146 | 0.1153 |
| CRC | 1,101 | 0.603 | 0.209 | 0.6093 |
| PCA | 1,340 | 0.081 | 0.092 | 0.0838 |

**Table 3.**
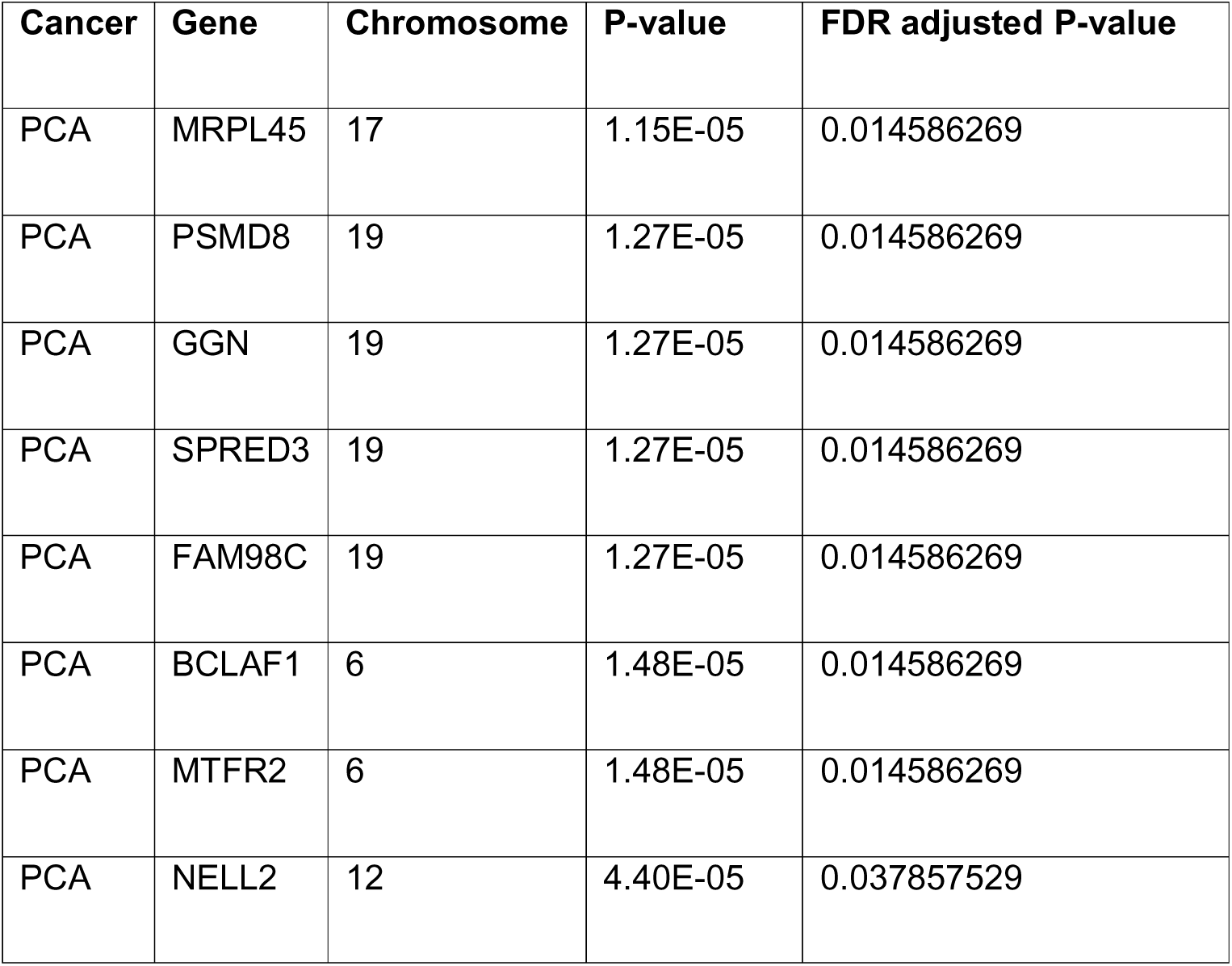
Prostate cancer gene-based burden test results at loci with FDR-significant associations.

GWAS identified five loci reaching genome-wide significance (p < 5×10−□), including four CRC loci and one novel BCa locus (Table 1).The strongest signals were observed for CRC, where several loci surpassed the conventional genome-wide threshold, including a variant near *RYR2* and a lead association on chromosome 22. Notably, *RYR2* has been increasingly implicated in various cancers through dysregulated calcium signaling, which may drive proliferative pathways and influence the tumor microenvironment ^40^. Fine-mapping of the chromosome 22 locus using African-ancestry LD reference data resolved a single, narrow credible set in which the index variant (rs117100284) carried the highest posterior inclusion probability. This high resolution reflects the power of African genomes, characterized by shorter LD blocks, to pinpoint causal variants more precisely than European cohorts, where long-range LD often obscures the true causal signal ^41^.

REGENIE gene-based GWAS revealed clear trait-specific patterns. While breast and colorectal cancer showed no genes surpassing FDR-adjusted significance, prostate cancer exhibited eight FDR-significant genes (q < 0.05), including *MRPL45, PSMD8, GGN, SPRED3, and BCLAF1*, clustering within biologically plausible regions on chromosomes 19q13 and 6q23. Transcriptome-wide association analysis yielded more limited discovery. Using African-ancestry GWAS summary statistics, ancestry-matched LD and GTEx v8 tissue weights, we identified *CYTH2* (TWAS Z = 4.15, p = 3.3×10−□) the only significant after Bonferroni correction in the PCa cohort. *CYTH2* encodes a cytohesin family member involved in ARF GTPase signalling and membrane trafficking and has emerging roles in cell migration and oncogenic signalling, making it a plausible mediator of prostate carcinogenesis. Notably, no significant TWAS signals were observed for BCa or CRC cohorts. This is not unexpected given that we restricted analyses to directly genotyped SNPs, and that GTEx expression models are trained predominantly in European-ancestry donors ^42,43^. Our results therefore underscore the urgent need for large-scale transcriptomic resources from African and African-diaspora populations.

Polygenic risk score performance varied substantially across cancers. For prostate cancer, ancestry-aware PRS demonstrated strong discrimination, with PRS-CSx achieving an odds ratio per standard deviation of 1.40 (95% CI: 1.20–1.63), an AUC of 0.89, and an incremental Nagelkerke ΔR² of 0.013 beyond age and sex. These results are consistent with prior evidence that prostate cancer PRS trained with African LD and multi-ancestry discovery data outperform direct transfer of European-derived scores (Conti et al., 2017; Ge et al., 2019). In contrast, PRS performance for colorectal and breast cancer was modest (AUC ≈ 0.63 and ≈ 0.77, respectively), with limited additional variance explained (ΔR² ≤ 0.006), reflecting both trait-specific genetic architecture and reduced portability of PRS across ancestries (Martin et al., 2019).

Using the cancer cohorts for validation, we benchmarked African-tuned PRS-CSx ^39^ scores against pre-existing scores from the PGS Catalog that were derived mainly from European-ancestry discovery GWAS. PRS performance varied substantially across cancers. For PCa, ancestry-aware PRS demonstrated strong discrimination, with PRS-CSx achieving an odds ratio per standard deviation of 1.40 (95% CI: 1.20–1.63), an AUC of 0.89, and an incremental Nagelkerke ΔR² of 0.013 beyond age and sex. These results are consistent with prior evidence that prostate cancer PRS trained with African LD and multi-ancestry discovery data outperform direct transfer of European-derived scores ^44^. PRS performance for CRC ^45^ and BCa ^46,47^ was modest (AUC ≈ 0.63 and ≈ 0.77, respectively) although close to metrics in previous literature, with limited additional variance explained (ΔR² ≤ 0.006). This reflects both trait-specific genetic architecture and reduced portability of PRS across ancestries ^48^. These findings align with recent landmark trans-ancestry meta-analyses, which demonstrate that while many risk loci are shared across populations, the refinement of variant weights using ancestry-specific LD is critical for achieving clinical-grade risk stratification in AA populations ^40,49^.

Collectively, our findings demonstrate that African-ancestry genomic analyses can reveal distinct heritability patterns, refine causal loci with higher resolution, and improve risk prediction when ancestry-aware models are applied. This study is limited by modest sample sizes for African-ancestry cancer cohorts in the UK biobank, which reduce power for GWAS, gene-based testing, and TWAS, and by reliance on directly genotyped variants that restrict cis-regulatory SNP coverage. The use of predominantly European-trained transcriptomic reference panels likely attenuated TWAS discovery despite African ancestry-matched LD and GWAS inputs. Future efforts should prioritize large-scale African-ancestry GWAS, sequencing-based variant discovery, and the development of African-centered transcriptomic and regulatory atlases. Such advances will be essential for improving causal inference, enhancing polygenic risk prediction, and ensuring that precision oncology benefits populations historically underrepresented in genomic research.

## Methods

### Data sourcing

This study was conducted under ethical approval from the Covenant Health Research Ethics Committee (CHREC), Covenant University, Ota, Nigeria (US DHHS IORG0010037; CHREC NHREC Registration Number: NHREC/CU-HREC/1/01/2025). Access to UK Biobank data was granted under UK Biobank Application ID 1026225.

Participants were included if their UK Biobank ^50^ ethnic background record indicated membership in one of the following categories corresponding to African or African-admixed backgrounds: African, Black or Black British, White and Black African,White and Black Caribbean & Any other Black background, leading to 9,091 individuals. Cancer outcomes were defined exclusively using Hospital inpatient diagnosis records based on ICD-10 codes such that cancer cases were identified using codes for malignant neoplasm of breast, prostate and colon, rectosigmoid junction, and rectum. Controls were sampled from individuals without the relevant ICD-10 cancer code recorded in inpatient diagnoses.

We obtained final set for Breast cancer (BCa) with 1565 individuals (166 cases, 1399 controls), Colorectal cancer (CRC), 1101 individuals (112 cases, 989 controls) and Prostate cancer (PCA) with 2511 individuals (250 cases, 2261 controls). Inputs were used directly to construct the African-ancestry analytic cohort for downstream GWAS, heritability estimation, PRS analyses, and TWAS.

### Quality Control

Genome-wide array genotypes were processed using the PLINK ^51^ software applying uniform sample and variant quality control criteria ^52^, including per-sample and per-variant missingness ≤1%, Hardy–Weinberg equilibrium filtering in controls (p < 1×10⁻□), and trait-specific minor allele frequency thresholds of minor allele frequency (MAF) ≥ 0.0005 for GWAS and PRS analyses and MAF ≥ 0.01 for SNP-heritability estimation (Supplementary Text 1 and 7.1).

### SNP-based heritability estimation (GCTA and FaST-LMM)

We estimated SNP-based heritability (*h*^2^) GCTA-GREML ^32^ and FaST-LMM ^33^, applied to the quality controlled autosomal SNP sets (Supplementary Text 2 and 7.2). We first constructed a genetic relationship matrix (GRM) from autosomal variants using GCTA, and then fitted a univariate GREML model for each cancer:

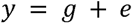

where (y) is the binary case–control phenotype (coded 0/1), 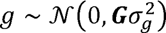 is the polygenic effect captured by the GRM (c), and 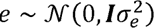 is the residual. GCTA reports genetic variance 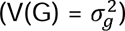, residual variance 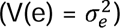 and total phenotypic variance (*V_p_* = V(G) + V(e)).

Observed-scale SNP heritability was defined as:

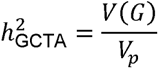

As a robustness check, we also estimated SNP heritability using FaST-LMM, which fits a linear mixed model with a GRM constructed from the same data. The FaST-LMM estimate,

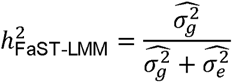

These values were compared to the GCTA estimates to assess sensitivity to software and modeling assumptions.

### Single-variant GWAS

We performed genome-wide association analyses using FaST-LMM ^33^ on the quality-controlled genotype datasets. For each cancer phenotype, we fit a linear mixed model of the form:

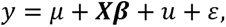

where *y* is the 0/1 case–control phenotype, *x* contains covariates (intercept, age at recruitment and sex), 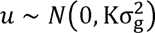 is the random polygenic effect with ***K***, Genomic relationship matrix, and 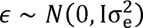 is the residual. FaST-LMM produced per-SNP association statistics (effect size, standard error, p-value) used downstream for Fine-mapping, TWAS (FUSION), and PRS-CSx.

### Gene-based association testing with REGENIE

We applied REGENIE ^53^ to perform gene-based association tests that aggregate multiple variants within protein-coding genes. For whole-genome prediction we ran REGENIE step 1 using ridge regression (stacked blocks of SNPs) to fit a whole-genome prediction model and obtain leave-one-chromosome-out (LOCO) predictions (Supplementary Text 7.3).

We then grouped variants into genes based on Ensembl/GENCODE ^54^ annotations (genomic coordinates in GRCh38). We evaluated the burden test and the variance-component test implemented in REGENIE using the LOCO predictions and adjusting for age and sex. For each gene and trait, REGENIE provided a gene-level effect estimate and p-value. We controlled for multiple testing using the Benjamini–Hochberg false discovery rate (FDR) procedure ^55^ applied to all gene-level p-values within each trait.

### Fine mapping of significant loci

We performed Bayesian fine-mapping of African-ancestry GWAS loci using the *sum of single effects* (SuSiE) regression framework ^38^. For each lead SNP, we defined a ±500 kb locus and harmonized GWAS effect estimates 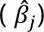 and standard errors (*SE_j_*) to the African-ancestry LD reference panel ^56^ (1000 Genomes AFR, GRCh38). Variants were required to match in genomic position and allele orientation. Local ancestry-matched LD matrices (*R*) were computed for all SNPs within each window.

SuSiE-RSS models the observed association statistics according to

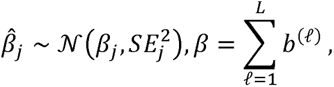

where each single-effect vector *b*^(ℓ)^ captures one putative causal signal, *L* is the maximum number of independent causal effects (signals) The model outputs variant-level posterior inclusion probabilities (PIP) and identifies 95% credible sets (CS) satisfying

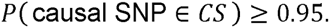

Fine mapping was conducted with *L* = 10 to allow for potential multi-signal loci (Supplementary Text 7.7).

### Transcriptome-wide association studies (TWAS) with FUSION

TWAS was conducted using the FUSION framework ^57^, which integrates GWAS summary statistics with genetically predicted gene expression. For each cancer phenotype, we used African-ancestry GWAS summary statistics generated in this study, ensuring that all downstream transcriptomic inference was driven by ancestry-matched genetic architecture. We combined our GWAS results with GTEx ^58^ v8 tissue-specific expression prediction models, using prostate (GTEx Prostate), breast (Breast Mammary Tissue), and colon (Colon Transverse) weights.

Because LD structure strongly influences TWAS effect estimation ^43^, all analyses used African-ancestry LD reference panels from the 1000 Genomes Project (GRCh38), ensuring ancestry-matched covariance estimates. For a given gene *g*, TWAS evaluates whether the genetically predicted expression:

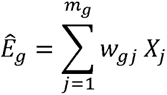

(where *w_gj_*is the cis-eQTL weight for SNP *j* and *x_j_* is the SNP genotype) is associated with disease. FUSION computes the gene-level Z-score as:

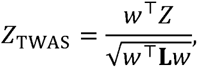

Where *z* is the vector of GWAS summary statistic SNP Z-scores, **L** is the ancestry-matched LD covariance matrix, *w* is the vector of tissue-specific prediction weights. TWAS was run separately for each chromosome and each cancer phenotype, producing a gene-level association statistic and p-value for all genes with available GTEx models and sufficient SNP overlap with the African LD panel (Supplementary Text 7.4).

### Polygenic risk score construction using PRS-CSx and PGS Catalog models

To derive ancestry-aware polygenic scores, we applied PRS-CSx ^39^, a Bayesian regression framework that leverages continuous shrinkage priors and ancestry-specific LD patterns. Discovery GWAS summary statistics for prostate, colorectal, and breast cancer were obtained from the GWAS Catalog ^59^ and represented large multi-ancestry meta-analyses: prostate cancer (GCST90479803; 61,885 cases and 511,807 controls), colorectal cancer (GCST90479781; 12,064 cases and 615,447 controls), and female breast cancer (GCST90479801; 3,125 cases and 52,184 controls).

Summary statistics were harmonized to GRCh38, allele-aligned, and filtered to variants shared with the African ancestry LD reference. PRS-CSx was run in single-population mode using the African ancestry panel, with default hyperparameters (a = 1, b = 0.5) and automatic global shrinkage estimation (φ = auto). Posterior mean effect sizes were generated chromosome-wise and used as SNP weights for PRS computation on the African ancestry.

To benchmark performance, we additionally evaluated pre-trained polygenic scores from the PGS Catalog^60^ that are based primarily on European-ancestry discovery datasets but remapped to GRCh38, including PGS000377 for colorectal cancer, PGS002242 for breast cancer, and PGS002241 for prostate cancer. Polygenic scores were computed in in our African ancestry cohort using *plink2*--score, generating PRS from either PRS-CSx posterior effect sizes or PGS Catalog weights (Supplementary Text 7.5 & 7.6).

### PRS evaluation on the African ancestry cancer cohorts

All PRS values were standardized within the target cohort using

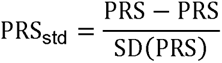

Where *PRS* is the raw polygenic score for, PRS is the sample mean and *SD*(PRS)is the sample standard deviation.

Covariate information, including age at recruitment and sex, was extracted from UK Biobank merged with genotype-based phenotype. All PRS, and prediction models were adjusted for age and sex.

For each cancer and scoring strategy (PRS-CSx or PGS), we fit a logistic regression model^61^:

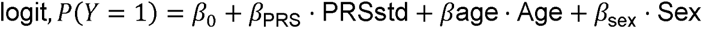

Odds ratios per SD and 95% confidence intervals were derived from the logistic regression coefficient for the standardized PRS (OR = *e^β^*^PRS^) following standard practice in logistic regression, while the full model (PRS + age + sex) discrimination was assessed using the area under the ROC curve (AUC). Pseudo-*R*^2^ was quantified using Nagelkerke’s *R*^2^, obtained by rescaling the Cox–Snell likelihood-based *R*^2^, and the incremental variance explained by the PRS was defined as 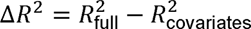^62^.

## Supporting information

supplmentary documentation

## Data and Code Availability

All analyses in this study were performed using publicly accessible resources or software listed in Supplementary Text 5 (Software and Versions). UK Biobank genotype and phenotype data were used under Application ID 1026225 and are available to approved researchers via the UK Biobank data access framework. GTEx v8 expression reference models were obtained from the GTEx Portal (https://gtexportal.org). GWAS summary statistics used for PRS-CSx benchmarking were obtained from the GWAS Catalog (accessions GCST90479803, GCST90479781, and GCST90479801).

## Software versions

All computational analyses were performed using Python 3.10.11 with package management via pip (pip3 for Python 3.10). Core Python libraries included NumPy v1.26.4, pandas v2.3.1, SciPy v1.15.3, statsmodels v0.14.5, scikit-learn v1.7.1, and matplotlib v3.10.3. Genome-wide association analyses using linear mixed models were conducted with FaST-LMM v0.6.12. Statistical analyses in R were performed using R v4.4.2 via Rscript, with key packages including susieR v0.14.2 for Bayesian fine-mapping and locuszoomr v0.3.8 for regional visualization. Transcriptome-wide association studies were performed using FUSION (Gusev et al.), accessed as external software from the official repository (https://github.com/gusevlab/fusion_twas), rather than as an R package installation.

Polygenic risk scores were constructed using PRS-CSx v1.1.0 (https://github.com/getian107/PRScsx/releases/tag/v1.1.0). Genotype data processing and quality control were performed with PLINK2 v2.00a6 (M1 build), SNP-based heritability estimation with GCTA v1.94.1 (gcta64), and gene-based association testing with REGENIE v4.0. All software versions were fixed throughout the study to ensure reproducibility, and full command-line execution details are provided in Supplementary Text 7 (Reproducibility commands).

## Author Contributions

Project conceptualization: D.E. and O.O.O., Computational data analysis: M.I. and D.E. &, Script and code development: D.E and M.I., Provision of data resources: O.O.O, Manuscript writing: D.E. M.I., C.C.E, O.E.E. and O.O.O, with approval from all coauthors. Project supervision: D.E. and O.O.O. Funding support: O.O.O.. All authors edited, reviewed, and approved the final version of the paper.

