## Supplementary material for "Deciphering the Genomic Architecture of Three Major Cancers in African-Ancestry Populations": supplmentary documentation

### Supplementary text

#### 1. Genotype data and quality control

Genome-wide array genotypes were processed using PLINK (v2.0/plink2) applying uniform sample and variant-quality control criteria. QC filters applied:

1. Per-sample missingness: <= 1%.
2. Per-variant missingness: <= 1%.
3. Hardy–Weinberg equilibrium in controls: P < 1e-6 (variants failing in controls were excluded).
4. Minor allele frequency thresholds: MAF >= 0.0005 for GWAS and PRS analyses; MAF >= 0.01 for SNP-heritability estimation using GCTA.
5. After QC the number of variants retained per trait: BRCA 526,048; PRC 509,066; CRC 515,671 (initial ~783,866 array markers).

#### 2. SNP-based heritability estimation (GCTA and FaST-LMM)

We estimated SNP-based heritability using both GCTA-GREML and FaST-LMM applied to autosomal variants passing QC (MAF >= 0.01 for heritability estimation).

GCTA pipeline (example commands):

1. Build GRM:

gcta64 --bfile uk_ba_qc_h2 --make-grm-bin --out uk_ba_qc_h2_grm

2. Create phenotype file (FID IID PHENO) and run REML:

\# write FID IID PHENO from .fam
awk '{print $1, $2, $6}' uk_ba_qc_h2.fam > uk_ba_qc_h2.pheno

gcta64 --grm uk_ba_qc_h2_grm --pheno uk_ba_qc_h2.pheno --thread-num 22 --reml --out uk_ba_qc_h2_reml

FaST-LMM: We also estimated h2 using FaST-LMM with a GRM constructed from the same variants; this served as a robustness check.

Reported heritability estimates (observed scale): BRCA GCTA h2 = 0.114 (SE = 0.146), FaST-LMM h2 = 0.115; CRC GCTA h2 = 0.603 (SE = 0.209), FaST-LMM h2 = 0.609; PRC GCTA h2 = 0.081 (SE = 0.092), FaST-LMM h2 = 0.084.

#### 3. TWAS (FUSION)

TWAS was conducted using FUSION: GWAS summary statistics from our African-ancestry analyses were combined with GTEx v8 tissue-specific prediction models (Prostate, Breast Mammary Tissue, Colon Transverse). Ancestry-matched LD covariance matrices (1000 Genomes AFR GRCh38) were used for TWAS computations. The TWAS Z-score for gene g is $Z_{T}WAS=\left( w^{T}Z \right)/sqrt\left( w^{T}Lw \right)$, where w are weights, Z are SNP z-scores, and L the LD covariance.

#### 4. PRS construction (PRS-CSx and PGS Catalog)

Discovery GWAS were obtained from GWAS Catalog: PRC (GCST90479803), CRC (GCST90479781), BRCA (GCST90479801). Summary statistics were harmonized to GRCh38 and aligned to the African LD reference. PRS-CSx was run in single-population mode with African LD reference (default hyperparameters unless stated). Posterior mean effect sizes were generated and used for scoring with plink2 --score. We also scored PGS Catalog models (PGS000377, PGS002241, PGS002242) after remapping to GRCh38.

PRS standardization and evaluation:

$$PRS_{std}=\left( PRS-mean\left( PRS \right) \right)/sd\left( PRS \right)$$

Logistic regression adjusted for age and sex was used to estimate OR per SD, 95% CI and p-value. AUC (full model) and Nagelkerke ΔR^2 (difference between full model and covariate-only model) were computed.

Visualization and QC

PCA of genotype data confirmed a single African/African-admixed cluster. QQ plots showed well-calibrated statistics (λ_GC: CRC=1.004, PRC=1.018, BRCA=0.998).

#### 5. Software and versions

Python: 3.10.11

pip: present (pip3 for Python 3.10)

Key Python packages:

numpy 1.26.4

pandas 2.3.1

statsmodels 0.14.5

scikit-learn 1.7.1

scipy 1.15.3

matplotlib 3.10.3

fastlmm 0.6.12

R / Rscript: R 4.4.2

susieR 0.14.2

locuszoomr 0.3.8

FUSION: external software at https://github.com/gusevlab/fusion_twas (not installed as an R package)

PRS-CSx: v1.1.0 (https://github.com/getian107/PRScsx/releases/tag/v1.1.0)

Genetics CLI tools:

PLINK2 v2.00a6 M1

GCTA v1.94.1 (gcta64)

REGENIE v4.0

#### 6. Compute resources / environment:

Local macOS: 8 CPU cores, UK Biobank RAP (DNA Nexus) cloud platform: (platform: https://platform.dnanexus.com/)

#### 7. Reproducibility commands (complete set)

This section provides the full set of commands used across the analyses. Adjust paths and thread counts as needed. The repository contains helper scripts referenced below (see file list in the repository root). Recommended environment setup (conda + R/Python packages) for the versions provided.

##### 1) Preprocessing and QC (PLINK2)

- Initial QC: remove samples/variants with >1% missingness
  plink2 --bfile /path/to/ukbiobank_raw_prefix \
   --geno 0.01 --mind 0.01 --make-bed --out uk_ba_qc_step1
- HWE filtering (controls only). If you have a controls list, use --keep
  plink2 --bfile uk_ba_qc_step1 --hwe 1e-6 --make-bed --out uk_ba_qc_step2
- MAF filter for GWAS/PRS (MAF >= 0.0005)
  plink2 --bfile uk_ba_qc_step2 --maf 0.0005 --make-bed --out uk_ba_qc_gwas_prs
- MAF filter for heritability estimation (MAF >= 0.01)
  plink2 --bfile uk_ba_qc_step2 --maf 0.01 --make-bed --out uk_ba_qc_h2

##### 2) GRM construction and GCTA REML

- Build GRM
  gcta64 --bfile uk_ba_qc_h2 --make-grm-bin --out uk_ba_qc_h2_grm
- Create .pheno file (FID IID PHENO) from .fam
  awk '{print $1, $2, $6}' uk_ba_qc_h2.fam > uk_ba_qc_h2.pheno
- REML (use available threads)
  gcta64 --grm uk_ba_qc_h2_grm --pheno uk_ba_qc_h2.pheno --thread-num 22 --reml --out uk_ba_qc_h2_reml

##### 3) REGENIE gene-based testing

- Step 1: whole-genome prediction (LOCO)
  regenie --step 1 --bed uk_gwas_prs--phenoFile pheno.txt --covarFile covar.txt --bsize 1000 --out regenie_step1
- Step 2: association using predictions
  regenie --step 2 --bed uk_gwas_prs--phenoFile pheno.txt --covarFile covar.txt --pred regenie_step1.pred --out regenie_step2

##### 4) TWAS with FUSION

- Prepare sumstats for FUSION: <https://github.com/gusevlab/fusion_twas>
- Run FUSION association for each gene/weight file (example)
  # Use fusion.assoc_test.R included in FUSION package
- Rscript fusion.assoc_test.R --sumstats fastlmm_chr4_fusion.sumstats --weights GTEx_Prostate.weights.gz --ref_ld_chr /path/to/AFR_ld/chr4 --out fusion_prostate_chr4.dat

##### 5) PRS-CSx and scoring

- Run PRS-CSx per-chromosome (example invocation; adapt to your PRS-CSx install)
  python PRSCSx.py --ref_dir /path/to/AFR_ld --bim_prefix /path/to/uk_gwas_prs--sst_file discovery_prscsx_sumstats.txt --out_dir prscsx_out --chrom 4 --phi auto
- Combine posterior betas into a single score file then compute PRS with plink2
  plink2 --bfile uk_gwas_prs--score prscsx_combined_weights.txt 1 2 3 header-read --out prscsx_scores

##### 6) Scoring PGS Catalog models with PLINK2

- Format PGS file for PLINK: columns SNP A1 weight (and optional header)
  plink2 --bfile uk_gwas_prs--score PGS002242_for_plink.txt 1 2 3 header-read --out pgs_PGS002242_prs

##### 7) Fine-mapping with SuSiE (example R script)

- Run SuSiE in R
- Rscript susie_finemapR.R --z locus_chr4_1603-1605/z.txt --ld locus_chr4_1603-1605/ld_matrix.txt --N 1565 --out susie_chr4_results

##### 8) Locus plotting (locuszoomr)

- Rscript make_locuszoom_plot.R --sumstats /path/to/uk_ba_chr4_sumstats.tsv --out locus_chr4.png --chr 4 --start 160300000 --end 160500000 --chrom-col Chr --pos-col ChrPos --p-col PValue --plot-type ggplot --width 10 --height 6 --dpi 600

### Supplementary figures

#### 1. Population structure plots across three cohorts

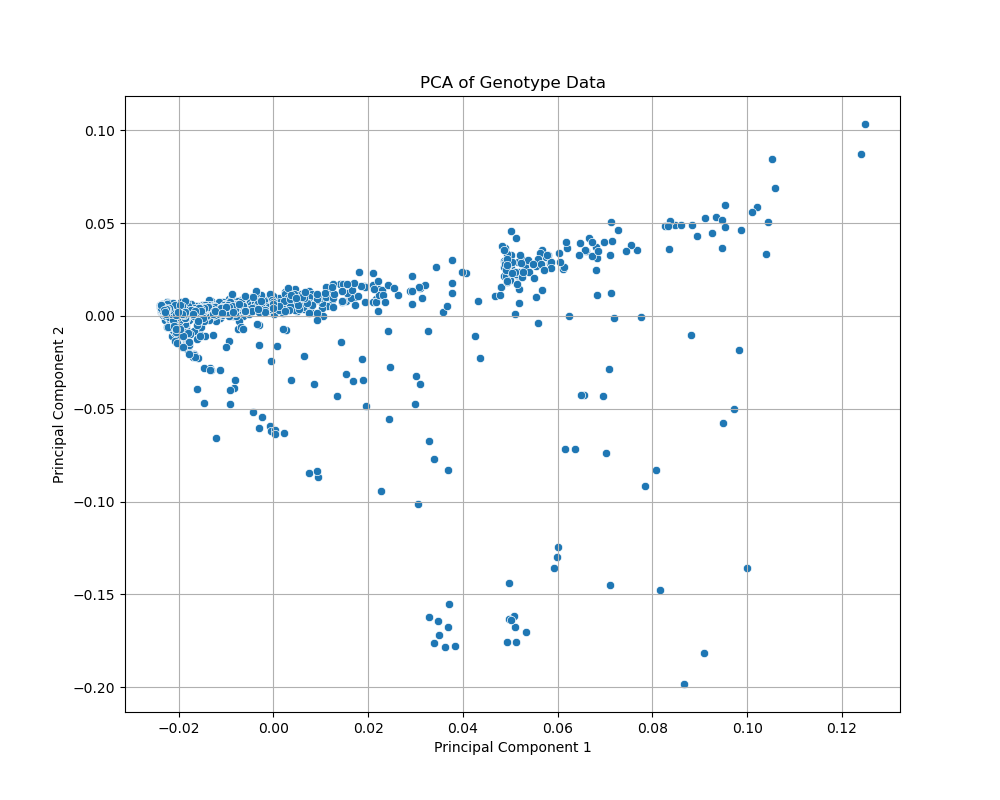

Figure 1 **Principal component analysis of African-ancestry cancer cohorts.** Scatterplots of the first two genotype principal components after QC in the African-ancestry subsets of UK Biobank, shown for colorectal cancer.

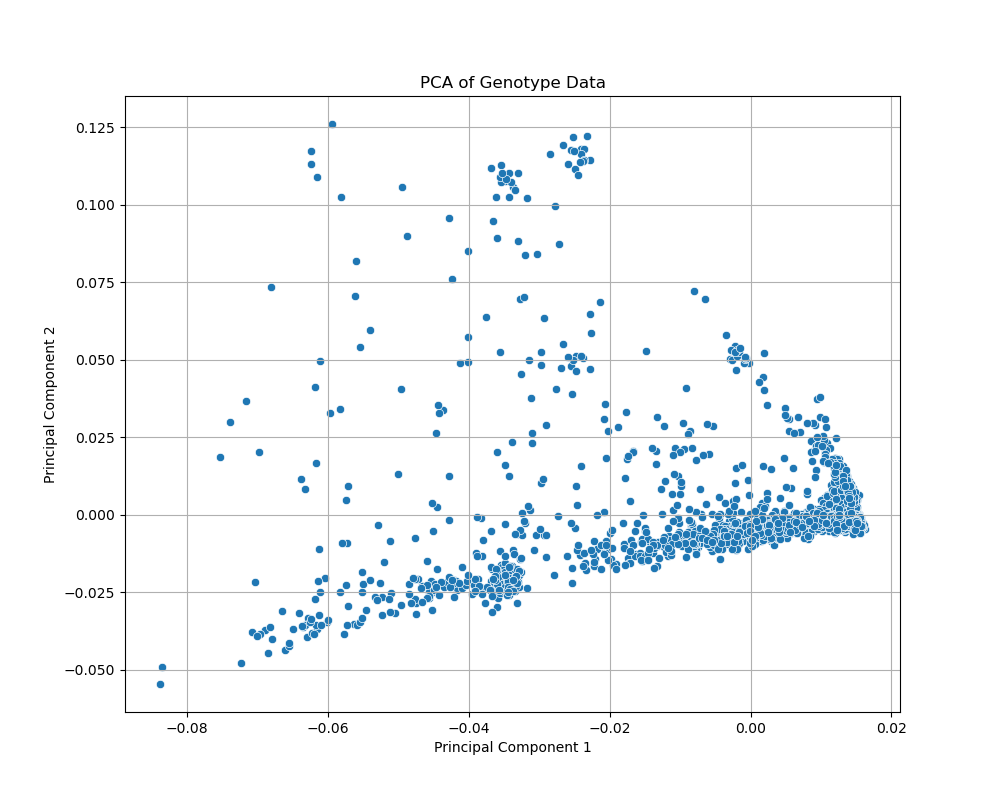

Figure 2 **Principal component analysis of African-ancestry cancer cohorts.** Scatterplots of the first two genotype principal components after QC in the African-ancestry subsets of UK Biobank, shown for prostate cancer

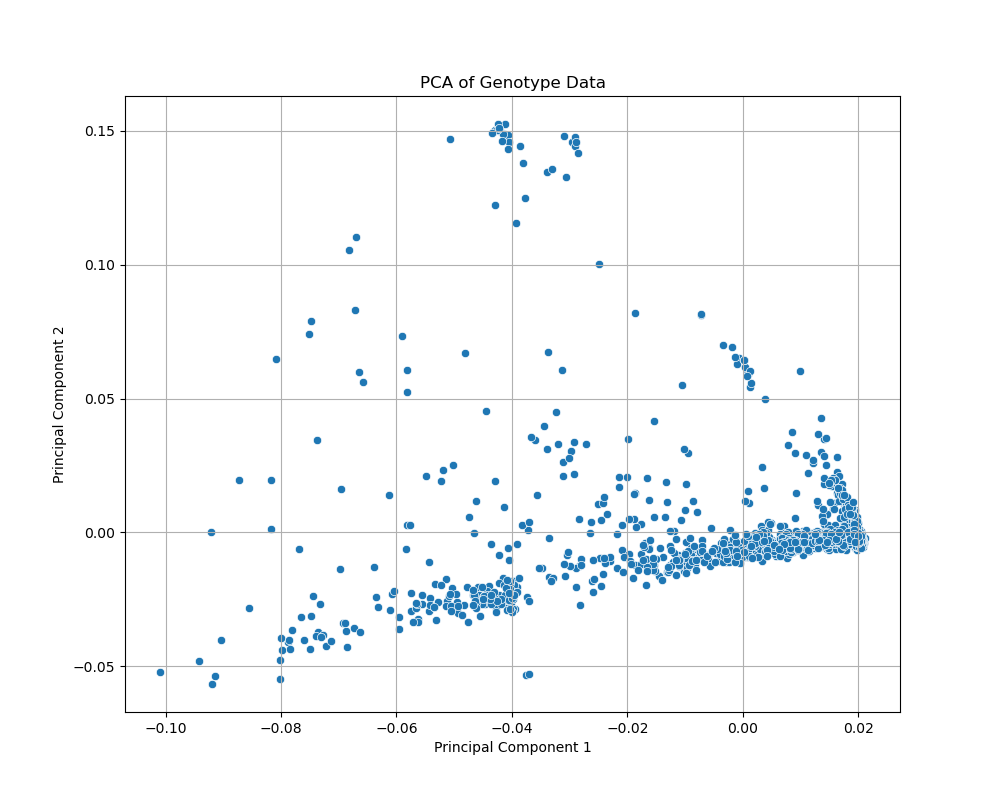

Figure 3 **Principal component analysis of African-ancestry cancer cohorts.** Scatterplots of the first two genotype principal components after QC in the African-ancestry subsets of UK Biobank, shown for breast cancer

#### 2. GWAS Lambda inflation factor across three GWAS done

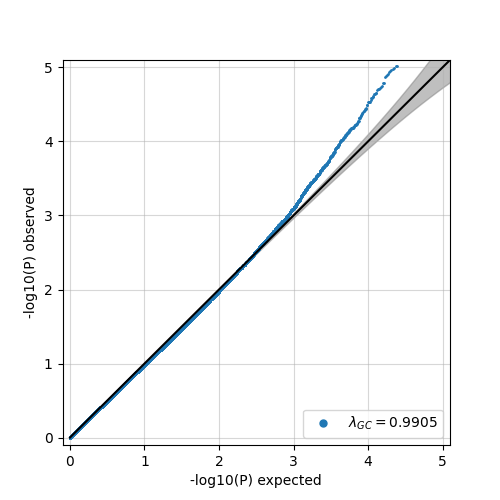

Figure 4 Quantile–quantile plot of FASTLMM GWAS p-values for breast cancer in African-ancestry UK Biobank participants. Observed versus expected −log₁₀(P) values are shown with a 95% null confidence band; the genomic inflation factor is λ_GC = 0.9905, indicating no evidence of systematic inflation.

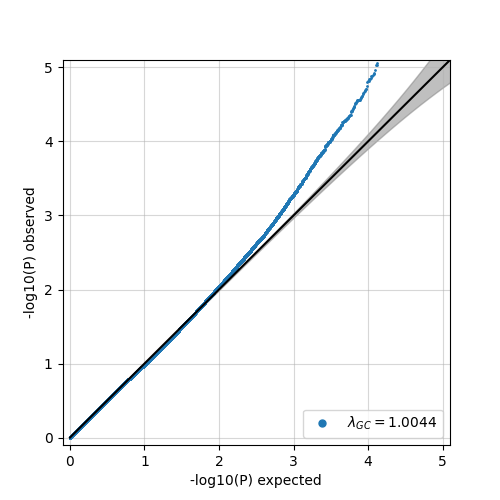

Figure 5 Quantile–quantile plot of FASTLMM GWAS p-values for colorectal cancer. The near-diagonal fit of observed versus expected −log₁₀(P) values (λ_GC = 1.0044) demonstrates well-calibrated test statistics with minimal inflation.

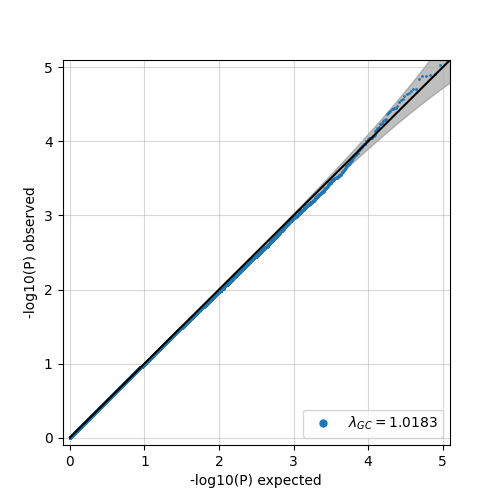

Figure 6 Quantile–quantile plot of FASTLMM GWAS p-values for prostate cancer. Observed −log₁₀(P) values closely follow the null expectation (λ_GC = 1.0183), supporting adequate control of population structure and relatedness.

#### 3. TWAS locus plots across for significant prostate cancer associations

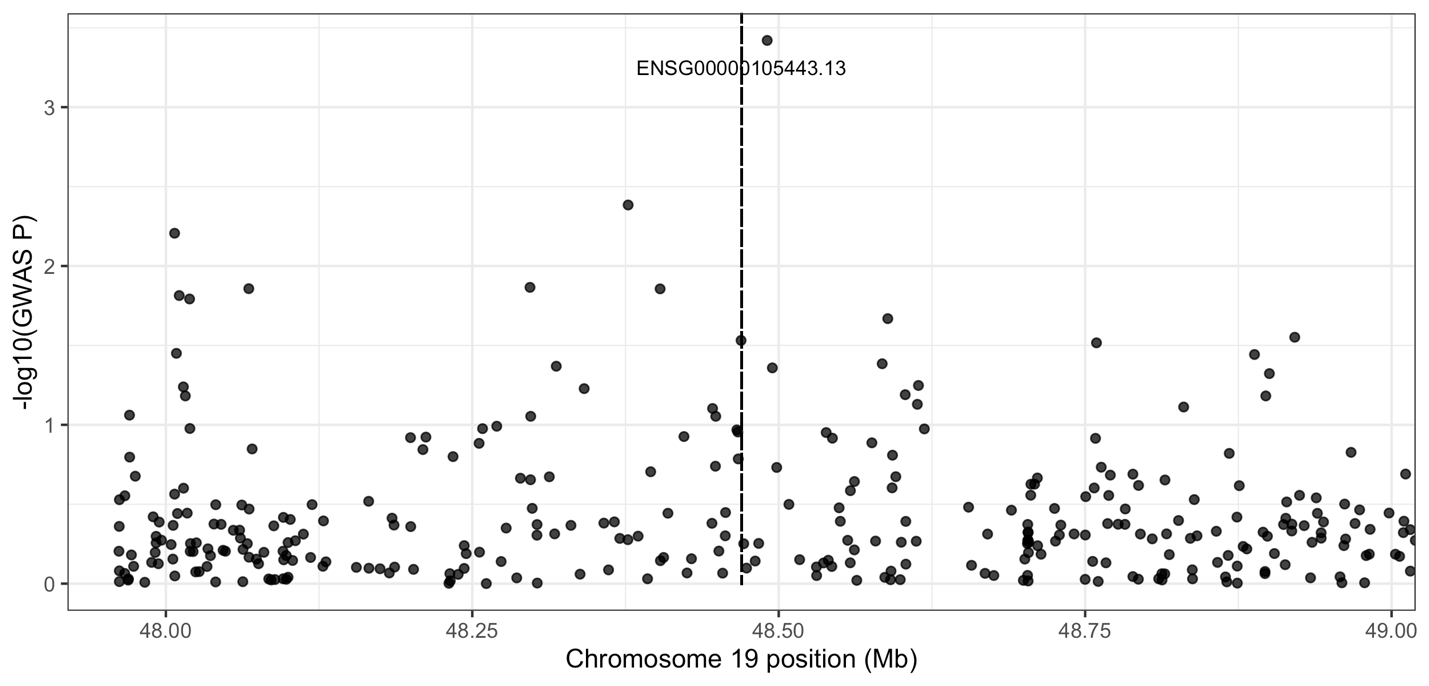

Figure 7 LocusZoom-style view of SNP associations across the 19q locus, with points coloured by local linkage disequilibrium (LD) with the lead variant in the region. The upper panel shows the LD-annotated SNP –log₁₀(P) values, and the lower panel shows the positions of nearby genes.

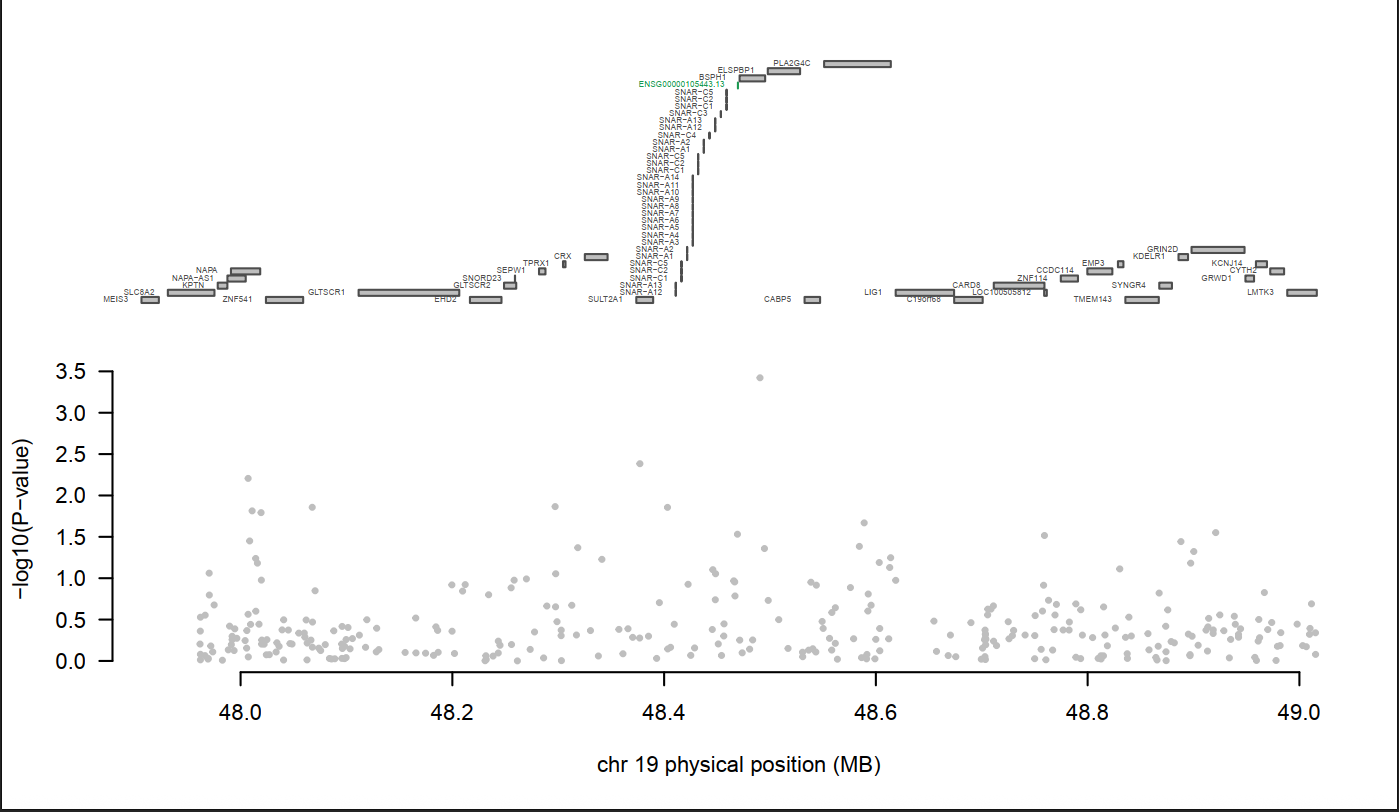

Figure 8 **Regional association signal at the chromosome 19 prostate cancer locus.**Focused regional GWAS plot for the same locus, highlighting the TWAS-implicated gene ENSG00000105443.13 (CYTH2) at ~48.5 Mb on chromosome 19 (vertical dashed line).

### Supplementary Tables

#### 1. Performance of polygenic risk scores for three cancers in African-ancestry UK Biobank participants.

Supplementary Table 1 Performance of polygenic risk scores for three cancers in African-ancestry UK Biobank participants. Logistic regression results for colorectal cancer (CRC), prostate cancer (PRC) and breast cancer (BRCA) using either PRS-CSx scores trained with an African-ancestry reference (PRS-CSx (AFR)) or trait-matched polygenic scores from the PGS Catalog (PGS000377, PGS002241, PGS002242). For each trait and PRS source, the table reports the number of cases and controls, the odds ratio (OR) per 1 standard deviation (SD) increase in PRS with 95% confidence interval (CI), p-value for the PRS term, area under the ROC curve (AUC) of the full model (age + sex + PRS), and Nagelkerke’s R²

| **Trait** | **PRS source** | **N** | **Cases** | **Controls** | **OR per SD (95% CI)** | **p-value** | **AUC** | **Nagelkerke R² (full)** | **Nagelkerke R² (covariates only)** | **ΔR² (PRS)** |
| --- | --- | --- | --- | --- | --- | --- | --- | --- | --- | --- |
| CRC | PRS-CSx (AFR) | 1101 | 112 | 989 | 1.186 (0.963–1.462) | 1.09×10⁻¹ | 0.63 | 0.0365 | 0.0314 | 0.0051 |
| CRC | PGS000377 | 1101 | 112 | 989 | 1.169 (0.961–1.422) | 1.18×10⁻¹ | 0.625 | 0.0359 | 0.0314 | 0.0045 |
| PRC | PRS-CSx (AFR) | 2511 | 250 | 2261 | 1.397 (1.198–1.629) | 1.95×10⁻⁵ | 0.888 | 0.3943 | 0.3812 | 0.0131 |
| PRC | PGS002241 | 2511 | 250 | 2261 | 1.297 (1.110–1.516) | 1.07×10⁻³ | 0.886 | 0.3947 | 0.3872 | 0.0075 |
| BRCA | PRS-CSx (AFR) | 1565 | 166 | 1399 | 1.112 (0.937–1.318) | 2.24×10⁻¹ | 0.771 | 0.2222 | 0.2204 | 0.0018 |
| BRCA | PGS002242 | 1565 | 166 | 1399 | 1.212 (1.021–1.439) | 2.81×10⁻² | 0.773 | 0.2344 | 0.2287 | 0.0057 |

#### 2. Performance of REGENIE gene based testing in African ancestry cohorts

Table 2 Top gene-based association signals for breast (BRCA), colorectal (CRC) and prostate (PRC) cancer in African-ancestry participants. Shown are results from additive burden tests (ADD) and SKAT-type tests (ADD-SKAT) under two variant inclusion masks

| Cancer | test | mask | Gene | top_pvalue | top_LOG10P | CHROM | GENPOS |
| --- | --- | --- | --- | --- | --- | --- | --- |
| BRCA | ADD-SKAT | Mask1.all | RIPK2 | 0.00231238 | 2.63594 | 8 | 89257806 |
| BRCA | ADD | Mask1.all | C12orf76 | 0.00164661 | 2.78341 | 12 | 109527028 |
| BRCA | ADD | Mask1.0.01 | NAT8 | 0.00013838 | 3.85893 | 2 | 73140723 |
| CRC | ADD-SKAT | Mask1.all | SRARP | 0.0017825 | 2.74897 | 1 | 15504236 |
| CRC | ADD | Mask1.all | MPI | 0.00056731 | 3.24618 | 15 | 74390005 |
| CRC | ADD | Mask1.0.01 | CTNNA2 | 9.24E-05 | 4.03422 | 2 | 78685231 |
| PRC | ADD-SKAT | Mask1.all | CHMP4B | 0.00029094 | 3.53619 | 20 | 33311348 |
| PRC | ADD | Mask1.all | MRPL45 | 1.15E-05 | 4.93752 | 17 | 37797023 |
| PRC | ADD | Mask1.0.01 | BMAL2 | 6.29E-05 | 4.20114 | 12 | 26832836 |
